# Voluntary Cyclical Distancing: A potential alternative to constant level mandatory social distancing, relying on an ‘infection weather report’

**DOI:** 10.1101/2020.05.02.20084947

**Authors:** Daniel Goldman

## Abstract

COVID-19 has significantly changed our daily lives. Stay-at-home orders and forced closings of all non-essential businesses has had a significant impact on our economy. While it is important to ensure that the healthcare system is not overwhelmed, there are many questions that remain about the efficacy of extreme social distancing, and whether there are alternatives to mandatory lockdowns. This paper analyzes the utility of various levels of social distancing, and suggests an alternative approach using voluntary distancing informed by an infectious load index or ‘infection weather report.’

## 2 Introduction

The outbreak of SARS-CoV-2 has caused a lot of changes in our daily lives. When policy makers around the world realized the threat of COVID-19, they began crafting guidance, and eventually started issuing stay-at-home orders. These orders have created significant economic disruption, and disruption to peoples’ lives. And there has been a question of how long they can be maintained. Already, lock-down orders are being lifted, at least in part, in some locations. But the question remains of what to do next, both in terms of dealing with the spread of SARS-CoV-2, and handling any future outbreaks of this nature.

Many of us are familiar with the catchphrase “flatten the curve.” The idea is that social distancing will reduce the rate at which the infection spreads, and thus reduce the burden on the healthcare system. However, these lockdown orders cannot be maintained indefinitely. One has to wonder how a premature end to a lockdown might impact the spread of the infection, and if there are any alternatives to such measures, and whether they might be more effective.

This paper has two goals. The first goal is to compare a number of modified SEIR models, in order to identify possible outcomes associated with the current lockdown efforts. The second goal is to identify potential ways to improve efforts to reduce the spread of both SARS-CoV-2, and infectious disease overall. The models in this paper rely on empirically estimated parameters, and take into account a number of factors, including social distancing, stratification of risk groups and hospital capacity. It then compares models in which a constant level of social distancing for a fixed period of time with modulated social distancing based on voluntary activity informed by disease surveillance.

## 3 Model Development

The core of these analysis is the SEIR compartmental model. Many alterations to the basic model have been made. There are two copies of each compartment, one for low risk individuals such as young people with minimal comorbidities, and one for high risk individuals such as the elderly and people suffering from various diseases or are otherwise in significantly poor health. For simplification, the focus for risk was on age. Mortality rates were also considered.

### 3.1 Base

The core model for each set of compartments is defined as follows:

*Ṡ* = *ρβSI*
*Ė* = *ρβSI* − *αE*
*İ* = *αE* − *γI* − *μI*
*Ṙ* = *γI*
*Ṁ* = *μI*
*N* = *S* + *E* + *I* + *R* + *M*

Here *β* is the product of the contact and transmission rates. *α* is the reciprocal of the incubation period, *γ* is the reciprocal of the clearing period post onset of symptoms, and *μ* is the mortality rate. And *ρ* is the adjustment to the contact rate, due to social distancing. Each of these cells are duplicated into a low risk and high risk set. For social distancing, it is assumed that there will be a greater amount of social distancing within the high risk population and between the low and high risk populations.

### 3.2 Data Sources and Estimates

#### 3.2.1 SEIR Parameters

Approximation of basic parameters comes from a number of sources. According to Peng et al. 2020, the latent time period, or the time it takes for a person to transition from exposed to infected, is approximately two days, giving *α* = 0.5[6]. The analysis also suggests that every contact is almost guaranteed to result in an infection: *β* ≈ 1. Because the model used in Peng et al. 2020 was complicated and did not calculate the unaided clearing of the infection, approximations from another source were used. D’Arienzo and Coniglio 2020 suggest that even in Italy where there is a significant COVID-19 burden, the basic reproduction number is between 2.43 and 3.10, which yields a range of 0.32 and 0.41 and for *γ*[3].

While *β* is approximated as 1, it is unlikely that the contact rates of individuals within the same age group is equal to the contact rate of individuals between the two groups. It is likely that within-group contact rate is higher than average, and that the between-group contact rates are lower than average. People within the high risk group are also more likely in general to maintain social distancing, and so this idea is also considered in approximating *β* for each type of interaction. Values were chosen s.t. the population weighted average summed to 1.

#### 3.2.2 Demographics

It was assumed that 84% of the population was in the low risk group and 16% was in the high risk group, and that there was one initial infection within each sub-population. This assumption is based on the fraction of the United States population aged 65 or older in 2018[7]. The United States population was slightly under 330M in 2018, so 330M was chosen for N[8].

#### 3.2.3 Hospital Capacity and Mortality Rates

Hospital bed capacity is estimated based on figures from COVIDACTNOW. The model assumes that there are roughly enough hospital beds for 0.22% of the population, with 60% capacity, and an emergency capacity build of roughly 200%[5]. As a conservative estimate, 0.1% was chosen for the capacity limit.

Infection and case fatality rates are highly dependent on a number of factors and vary based on the quality of the health care system, the age of the patient, and comorbidity. Mortality seems to be orders of magnitude higher in at risk populations compared to low risk populations. COVIDACTNOW estimates a case fatality rate of 1.1% with an additional 1% if hospitals are overburdened[5]. However, it does not stratify by risk group.

The base model starts with the assumption that the case fatality rate is 0.1% for low risk populations and 10% for high risk populations. Assuming that being over-capacity increases the risk of death among the low risk population by 50% and the high risk by 200%, that would yield a case fatality rate of 0.15% and 30% respectively. The 50% figure is still higher than the relative risk at 10 days, for general ER visits, but within the 95% CI of 1.04-1.72[9].

However, not every infection meets the criteria of being a case. There are many asymptomatic and subclinical infections for SARS-CoV-2. By one estimate, the number of infections was 50 to 85 times higher than the reported number of cases[1]. However, it’s quite possible that a number of those infections resulted in deaths that were not reported. Furthermore, since it is more likely that high risk individuals are more likely to show dangerous symptoms, and their status as being high risk yields a greater rate of testing, the mortality rates of the high risk group received a smaller adjustment. Low risk mortality rates were divided by 20 and high risk mortality rates were divided by 5.

### 3.3 Parameters

N = 330,000,000
*α* = 0.5
*β*_1_ = 1.075 - The adjusted contact-transmission rate within the healthy population
*β*_2_ = 0.75 - The adjusted contact-transmission rate between both populations
*β_3_* = 0.9 - The adjusted contact-transmission rate within the high risk population
*γ* = 0.37 - Average between the low and high bound estimates for the clearing rate
*ρ* = *[variable]* - The base social distancing coefficient
*ρ*_1_ = *ρ* - Social distancing coefficient for low risk group
*ρ*_2_ = 0.8*ρ* - Social distancing coefficient between the low and high risk groups
*ρ*_3_ = 0.8*ρ* - Social distancing coefficient for high risk group
*μ*_1_ = 0.0005 - Mortality rate of low risk group under optimal conditions
*μ*_2_ = 0.050 - Mortality rate of high risk group under optimal conditions
*μ*_3_ = - Mortality rate of low risk group under sub-optimal conditions
*μ*_1_ = - Mortality rate of high risk group under sub-optimal conditions
*κ* = 0.001 - Percent of population infected before hospitals are over capacity

## 4 Models and Model Analysis

A number of analyses were performed. First, it seemed useful to simulate how different levels of social distancing impacted the progression of the epidemic, when social distancing is maintained for a fixed period time period of sixty days, with initial onset 60 days after the first infection, representing lag between initial discovery of the disease and decision to engage in mandatory social distancing.

### 4.1 Relative Mortality Across Social Distancing Parameters

For social distancing parameters ranging from 0.1, representing extreme social distancing, to 1, representing no social distancing, a social distancing parameter of 0.65 appeared most effective, with an approximate reduction in mortality of 40%. A social distancing coefficient of less than 0.65 caused an uptick in mortality rates from that low. The minimum estimated mortality rate, for the entire progression of the epidemic was approximately 0.0062 or 620 per 100,000. For the United States, that would imply a final death toll of approximately 2 million under moderate social distancing, and 3.3 million without any social distancing.

**Figure 1:**
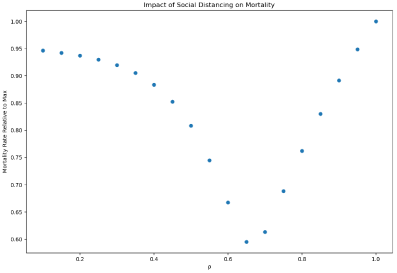
Mortality Relative to Base Mortality

Looking at the progression of the epidemic, for *ρ* = 0.4 helps to understand why values less than 0.7 result in higher mortality rates.

**Figure 2:**
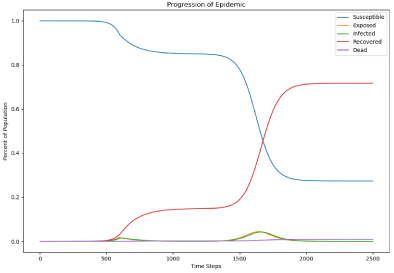
Epidemic Progression With *ρ* = 0.4

Each time step represents 0.1 days. At the start of the outbreak, there is a significant reduction in infections and deaths, but there is a spike in both shortly after the end of the social distancing effort. Rather than flattening the curve, the more extreme social distancing measures appear to delay the peak, allowing “pressure” to build up due to a high reserve of susceptible individuals. In order for more extreme continuous social distancing measures to be effective, they would therefore have to be maintained until an alternative, such as a vaccine, is produced.

### 4.2 Cyclical Distancing

As early as the beginning of April, it became apparent that a one time social distancing effort may be not be enough to cope with the COVID-19 epidemic[4]. However Kissler et al. 2020, while recognizing this issue, did not seek to establish a specific protocol for when social distancing should be engaged and disengaged[4]. Assuming that it is possible to obtain reasonably high resolution data for a region, the public health community should be able to put out daily and weekly advisories. These advisories can be used to promote voluntary social distancing, during periods of high levels of infectious load.

Because the social distancing measures would be temporary, and because there would be less uncertainty, because of the clear conditions for distancing recommendations, a slightly more extreme level of social distancing should be possible, so a *ρ* = 0.5 was chosen. A threshold of 0.0005 for the 7 day moving average of infections was chosen, because it was half the estimated maximum safe load that hospitals could handle. Furthermore, because reports of low infectious load could yield a false sense of safety, *ρ* = 1.05 was chosen during periods when social distancing was not engaged. The graphical results are detailed in the following figure.

**Figure 3:**
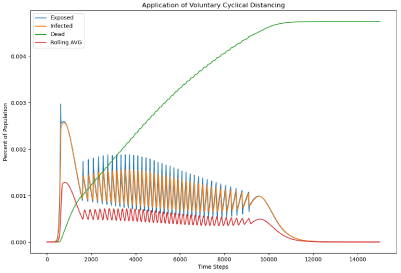
Application of Voluntary Cyclical Distancing

The results of the analysis are interesting. Voluntary cyclical social distancing, using the parameters chosen, results in a significantly extended curve. It takes around 1,200 days for the infection to fully burn itself out under this scenario. However, mortality rates are also much lower under this scenario, with the final mortality rate coming in at roughly 0.0047 or 470 per 100,000 people. In this model, there was no fixed time at which social distancing was expected to start. However, it took roughly 62 days for the infectious load to build enough to trigger the first distancing event, which is on par with the first model. All together, there were 39 periods of social distancing, with the last ending roughly 911 days after the initial infection.

## 5 Discussion

Feasibility depends on a number of factors, including the ability to collect sufficient data to generate infectious load indices for a desired geographic scale, and the ability to actually engage in social distancing on a voluntary basis. One question is whether we can collect enough information to create such an infectious disease index. While it would take a number of years to create a robust index that can be used in general cases, there should be no issue with creating an index specific to COVID-19. While it is true that a lot of countries, especially the United States, are unable to test anywhere close to every individual, random sampling can give us a significant amount of information on infectious load. Such random sampling requires a fraction of the number of tests that are needed to identify and isolate every infection.

With random sampling, integrated with other data gathering techniques, it is possible to have a fairly reasonable understanding of the progression of this epidemic. Much of the determination of cost effectiveness and ability to report will depend on the level of resolution we wish to have. If the goal is to have a composite state-wide infection index, fewer tests will be needed per day. Creating a county level index would be significantly more expensive.

By reporting information to the population, we can alter behavior so that voluntary social distancing can be modulated as infection dynamics change. This modulation while extending the duration of the infection, significantly flattens the curve, without mandatory stay-at-home orders. This flattening significantly extends the duration of the pandemic, but reduces the burden on the healthcare system and reduces the overall mortality rate. Additionally, given the level to which the epidemic period is lengthened, such measures would give time to produce treatments and prophylactics.

In this analysis, the initial length of time to bring the infection rate below threshold, and thus end the first social distancing event was slightly greater than 90 days, which is significantly longer than the 60 day social distancing measure used in the first simulation, and likely a bit longer than the length of time for which social distancing measures will be in place for COVID-19, at least during the initial wave. However, current lockdown measures are driving the social distancing coefficient, ρ, well below 0.5, which is far more extreme, and which cannot be maintained for as long a period of time.

Regarding the model which uses a single level of social distancing for a fixed period of time, it is concerning that the optimal social distancing coefficient is 0.65, which is likely far below the current level of social distancing, caused by the forced shutdown of all “non-essential” operations. While such extreme social distancing may be useful if limited to those within the at risk group, and between low and high risk groups, it does not seem appropriate for the general population.

There are a number of assumptions about parameter estimates that were made to test these models. In particular, the mortality rates for low and high risk populations are rough estimates. However, while they will alter the specific values in terms out fatality outcome, they should have little impact on the progression of the infection itself. Still, additional research into the case, and infection mortality rates, and greater stratification of risk levels would help give a better picture of potential outcomes and efficacy of existing and future solutions.

Improvements to this model however could be made. Additional stratification is possible, and empirical estimates of actual contact rates could be calculated with future research. Research by Chitnis et al. 2013 may be of interest, as it looks at a highly stratified population and uses empirical estimation of heterogeneous mixing between age groups[2]. Guesses in this area is likely to have some effect on the optimal social distancing level. But again, it should not impact the comparison between the fixed and cyclical distancing models to any significant extent.

While elements of this model are specific to SARS-CoV-2, the general dynamics would still apply to other infections. With a little bit of time and resources applied to the problem, a general reporting system for infectious disease could be implemented. This system could be used to help reduce the severity of flu seasons, and during future epidemics. Such infection weather reports, so to speak, could become part of the new normal. Moreover, this system could be especially useful if the COVID-19 epidemic enters a seasonal pattern due to limited generation of immunity, which is a concern that has been voiced[4].

Regarding the ability to actually engage in voluntary social distancing, a major concern is the ability to take off from work. If social distancing efforts needed to be extreme and extended for a long period of time, this issue would be more problematic. Given the reliance on our job, and the general inability to take off of work for extended periods of time to recover, this issue applies to situations outside of COVID-19 as well. Anyone who feels sick, especially if they have a fever, cough, or other symptoms of a potentially infectious disease, should engage in social distancing. However, financial needs often override wisdom and public safety guidelines.

However, given that simply reducing the average contact rate by 50% is enough to significantly reduce the rate of spread of the infection, a few minor decisions are all it would take. Moderately reducing frequency and lengths of outings, and being increasingly aware of one’s surroundings are all it would take to significantly reduce average contact rate. It is also likely that during periods where there are reports of high levels of infectious load, employers would be more willing to let an employee stay home and or cut back services.

Finally, research should be conducted into creating a composite index rather than trying to produce an index for a single pathogen. Analyzing the utility and efficacy of such a composite index will be far more complicated, as it would require the incorporation of numerous pathogens. However, because the public health system would not be seeking to limit a single infection but rather the bulk of infections, it might actually be easier to produce an effective index for an aggregate.

## Data Availability

The models used in this analysis are available through our github repository.

https://github.com/dgoldman0/socialdistancing_final

## References

[1] Eran Bendavid et al. “COVID-19 Antibody Seroprevalence in Santa Clara County, California”. In: *medRxiv* (2020). DOI: 10.1101/2020.04.14.20062463. eprint: https://www.medrxiv.org/content/early/2020/04/17/2020.04.14.20062463.full.pdf. URL: https://www.medrxiv.org/content/early/2020/04/17/2020.04.14.20062463.

[2] Nakul Chitnis, J. M. Hyman, and Sara Y. Del Valle. “Mathematical models of contact patterns between age groups for predicting the spread of infectious diseases”. In: Mathematical Biosciences and Engineering 10.5/6 (Aug. 2013), pp. 1475–1497. DOI: 10.3934/mbe.2013.10.1475. URL: https://doi.org/10.3934/mbe.2013.10.1475.

[3] Marco D’Arienzo and Angela Coniglio. “Assessment of the SARS-CoV-2 basic reproduction number, R0, based on the early phase of COVID-19 outbreak in Italy”. In: Biosafety and Health (Apr. 2020). DOI: 10.1016/j.bsheal.2020.03.004. URL: https://doi.org/10.1016/j.bsheal.2020.03.004.

[4] Stephen M. Kissler et al. “Projecting the transmission dynamics of SARS-CoV-2 through the postpandemic period”. In: Science (Apr. 2020), eabb5793. DOI: 10.1126/science.abb5793. URL: https://doi.org/10.1126/science.abb5793.

[5] Master CoVidActNow CoVid-19 Model - Google Drive. URL: https://docs.google.com/spreadsheets/d/1YEj4Vr6lG1jQ1R3LG6frijJYNynKcgTjzo2n0FsBwZA/htmlview# (visited on).

[6] Liangrong Peng et al. “Epidemic analysis of COVID-19 in China by dynamical modeling”. In: (Feb. 2020). DOI: 10.1101/2020.02.16.20023465. URL: https://doi.org/10.1101/2020.02.16.20023465.

[7] Population ages 65 and above for the United States (SPPOP65UPTOZSUSA) — FRED — St. Louis Fed. URL: https://fred.stlouisfed.org/series/SPPOP65UPTOZSUSA (visited on).

[8] Population ages 65 and above for the United States (SPPOP65UPTOZSUSA) — FRED — St. Louis Fed. URL: https://fred.stlouisfed.org/series/SPPOP65UPTOZSUSA (visited on).

[9] Drew B Richardson. “Increase in patient mortality at 10 days associated with emergency department overcrowding”. In: Medical Journal of Australia 184.5 (Mar. 2006), pp. 213–216. DOI: 10.5694/j.1326-5377.2006.tb00204.x. URL: https://doi.org/10.5694/j.1326-5377.2006.tb00204.x.

